# Subgroups of adult-onset diabetes: a prospective follow-up study of progression of insulin resistance and deficiency and association with liver steatosis and fibrosis

**DOI:** 10.64898/2025.12.01.25340818

**Authors:** Aleksi Laitinen, Annemari Käräjämäki, Aki J Käräjämäki, Samu Kurki, Liisa Hakaste, Olof Asplund, Johan Hultman, Kaj Lahti, Mikko Lehtovirta, Emma Ahlqvist, Tiinamaija Tuomi

**Affiliations:** Institute for Molecular Medicine Finland and Research Program of Clinical and Molecular Medicine, University of Helsinki, Helsinki, Finland; Department of Endocrinology, Vaasa Central Hospital, Wellbeing Services Area of Ostrobothnia, Vaasa, Finland; Department of Endocrinology, Turku University Hospital, Turku, Finland; Diabetes Unit of Ostrobothnia, Wellbeing Services County of Ostrobothnia, Vaasa, Finland; Department of Gastroenterology and Hepatology, Vaasa Central Hospital, Wellbeing Services of County of Ostrobothnia, Vaasa, Finland; Folkhälsan Research Center, Helsinki, Finland; Lund University, Malmö, Sweden; Helsinki University Hospital, Helsinki, Finland

**Keywords:** Cluster analysis, glucagon stimulation, liver steatosis, liver fibrosis, insulin resistance, insulin secretion

## Abstract

**AIMS:** To assess changes in insulin secretion and sensitivity in diabetes subgroups (from cluster analysis), the stability of the subgrouping, and the association between steatotic liver disease (SLD), liver fibrosis (LF) and tissue insulin resistance.

**METHODS:** Participants from a regional diabetes register were studied within three years from diagnosis and 2-10 years later with fasting laboratory tests (n=547). A subset (n=194) participated in an investigation with i.v. glucagon-insulin tolerance test, and elastography.

**RESULTS:** Differences between subgroups attenuated during the follow-up. Obesity- and age-related subgroups (MOD & MARD) were stable with >76% retaining the same cluster, whereas that hold for <25% in the groups characterized by insulin deficiency or resistance (SIDD & SIRD). Despite stable BMI, HOMA2-IR decreased significantly, but remained highest, in SIRD. SLD/LF was most prevalent in SIRD (70%/42%) and MOD (60%/41%); BMI mainly explained the subgroup differences. Independently of BMI or SLD, LF was associated with insulin resistance in liver (OR 3.90 [95% CI 2.05-7.42]) and adipose tissue (1.14 [1.06-1.22]) and insulin sensitivity in muscle (0.33 [0.17-0.64]).

**CONCLUSIONS:** Subgrouping should be performed near diagnosis, as the more severe phenotypic features attenuate with time (or treatment). To assess the risk of LF, the degree of insulin resistance should be evaluated, not only BMI.

## 1. INTRODUCTION

Adult-onset diabetes includes heterogenous disease phenotypes and metabolic dysfunctions, which translate only partially into different treatment or follow-up trajectories. In 2018, we suggested five subgroups based on presence or absence of GAD-antibodies (GADA) and a cluster-analysis of five clinical variables measured near diagnosis of diabetes: HbA1c, age, BMI and estimates of insulin secretion and -resistance at fasting (homeostasis model assessment 2, HOMA2-B and HOMA2-IR, respectively). [1]

Severe autoimmune diabetes (SAID), including GADA-positive type 1 diabetes and latent autoimmune diabetes in adults (LADA), and GADA-negative severe insulin-deficient diabetes (SIDD), have decreased insulin secretion and marked hyperglycemia. Conversely, individuals grouped as moderate obesity- (MOD) or age-related (MARD) diabetes along with severe insulin-resistant diabetes (SIRD) are diagnosed with only slightly elevated HbA1c levels.

The subgroups differ regarding the risk of comorbidities [1, 2]. In particular, the SIRD group has earlier and more prevalent kidney disease, and the SIDD and SIRD groups the highest age- and sex-adjusted risk of incident nephropathy and myocardial infarction, despite a different glycemic level. While the SIRD group also seems to have more steatotic liver disease (SLD) (based on recorded ICD-codes, ALT [2] and MRI imaging in a small study [3]), the association with liver fibrosis has been even less evaluated.[4] Insulin resistance is known to increase the risk of liver steatosis, fibrosis and cirrhosis, [5, 6] which are associated with cardiovascular events and mortality. [7, 8]

While these subgroups have been replicated in multiple cohorts, [9] only one study has looked at the progression of insulin deficiency and resistance over time [3], and little is known about the association with insulin resistance in different tissues. These aspects could aid in dissecting the pathophysiological factors contributing to the heterogeneity of diabetes [10], and inform targeted treatment and follow-up of diabetes [11] and liver comorbidities. [12–15]

In this prospective follow-up study, we studied progression of insulin deficiency and resistance as well as the stability of the cluster assignment. Moreover, we compared fasting and stimulated measures of insulin secretion and action, as well as the association between SLD, LF and insulin resistance in different tissues overall and within the subgroups of diabetes.

## 2. RESEARCH DESIGN AND METHODS

### 2.1 Study population

The Diabetes registry of Vaasa (DIREVA) is a Finnish regional study recruiting individuals with diabetes in the Vaasa Hospital District since 2007. At the end of 2023, of about 10 000 adults with diabetes in the region, 8187 with any type of diabetes had been registered (with signed consent) by diabetes nurses and provided blood samples [DIREVA lab-1: HbA1c, fasting plasma glucose (FPG) and serum/plasma C-peptide (S/P-Cpe)]. All procedures were performed in compliance with relevant laws and institutional guidelines. Ethics committees of Vaasa Hospital District, the Wellbeing Services County of Southwest Finland (VARHA/2406/13.02.02/2024[13.2.2024]) and the Hospital District of Southwest Finland (ETMK Dnro:48 /1801/2014[16.3.2021]) approved the study.

The follow-up study included 547 individuals (Figure S1): Group_A_, 500 with questionnaire data and new laboratory samples; Group_B_, 194 participating in a study visit (overlap between groups, N=147). The inclusion criteria were: 1) age at diagnosis of diabetes ≥18 years (and ≤80 years in Group_B_); 2) registration data within 3 years from diagnosis to assign the cluster subgroups [1] (BMI, age at diagnosis, and DIREVA lab-1); 3) time-difference of 2-10 (Group_A_) or 3-10 (Group_B_) years from the registration. We invited all individuals fulfilling the criteria for Group_A_, and all those belonging to the SAID, SIDD, and SIRD subgroups, and a random sample from the MOD and MARD subgroups for Group_B_. The cluster-based subgroups were assigned using the ANDIS cohort [1, 2] as a reference group.

All participants were sent a questionnaire (including, but not restricted to, questions on self-measured weight and waist circumference, medication, weight development, and alcohol consumption), and fasting blood samples were drawn for the same tests as at registration (DIREVA lab-1), as well as for serum alanine aminotransferase (ALT), aspartate aminotransferase (AST), glutamyl transferase (GT), creatinine, lipids, and blood hemoglobin (Hb), leukocytes and platelets (DIREVA lab-2).

### 2.2 Study visit (Group_B_)

A research nurse measured the weight, height, waist and hip circumference, heart rate, blood pressure, and fat free mass (Body composition analyzer BF-350, Tanita). Fasting (10-12 hours) venous blood samples were drawn for DIREVA lab-1/2, serum/plasma insulin, glucagon, GADA and IA-2A (Islet antigen-2 antibodies), followed by a combined i.v. glucagon and insulin tolerance test (GITT). [16] An i.v. bolus of 0.5 mg glucagon was administered at 0 min and of insulin aspart (0.05 IU/kg diluted to 10 IU/ml, NovoNordisk, Bagsværd, Denmark) at 40 min. Blood samples were drawn for PG at 0, 40, 45, 51 and 60 min, and for S-Cpe at 0 and 6 min. Insulin sensitivity was estimated using the first-order rate constant for glucose disappearance between 40 and 60 min (K_ITT_=ln 2/T_1/2_x100). [16]

The exclusion criteria for glucagon administration were FPG >10 mmol/l, previous measurement of S-Cpe <0.2 nmol/l, insulin pump therapy, systolic blood pressure >200 or diastolic >110 mmHg, or questionnaire replies indicating risk of heart-related adverse effects.

In case of insulin-treatment (30 individuals), the basal insulin was continued, but the morning dose of glargine 100 IU/ml, detemir or biphasic insulin aspart was postponed until after the tests; the insulin pump dosage was reduced by 20% one hour before the examination. Liraglutide and exenatide (N=5) were paused 2 days, semaglutide (N=23) one week, and oral glucocorticoids one month before the study. On the morning of the visit, participants did not take any medications besides anticoagulants.

We excluded individuals from some analyzes (Fig S1b) due to previous obesity surgery, liver metastasis, or being extreme outliers.

### 2.3 Liver fat content and liver fibrosis (Group_B_)

On a separate day (median [interquartile range, IQR] difference 28[37] days), vibration-controlled transient elastography (VCTE with M/XL probes) was performed after a 3-hour fast to analyze liver steatosis (continuous attenuation parameter, CAP) and fibrosis (liver stiffness measurement, LSM) [17] using FibroScan® 530 (Echosens, Paris, France) from right-sided central axis line with the patient at supine position (requirement: ten successful measurements and an IQR/median ratio <30%). We used 288 and 302 dB/m as cut-off values for CAP [17, 18] to qualitatively estimate SLD (Table S6).

### 2.4 Laboratory tests

The DIREVA lab-1/2 tests were analyzed by the Vaasa Central Hospital Laboratory or their subcontracters. B-HbA1c nearest to registration date was obtained from the laboratory database (median 0 [IQR 59] days, 80%/89% within 100/365 days of other measurements). GADA and IA-2A were analyzed using Elisa (RSR Limited, Cardiff, UK), and free fatty acids (FFA) using NEFA-HR(2) Assay (Fujifilm Wako chemicals Europe GmbH, Germany). Fasting adipose tissue insulin resistance index (Adipo-IR) was calculated as FFA (mmol/l) x insulin (μU/ml). Glucose was analyzed at the study centre (HemoCue Glucose 201+, HemoCue AB, Ängelholm, Sweden).

S/P-Cpe was analyzed with immunochemical assays (Advia Centaur XPT, Siemens Healthineers AG, Forchheim, Germany; Cobas e801 ECLIA, Roche Diagnostics, Mannheim, Germany). In our research laboratory, we also analyzed from stored (−20°C) S/P samples glucagon (ELISA 10-1271-01, Mercodia AB, Uppsala, Sweden), lipids (Thermo Indiko, Waltham, Massachusetts, USA), and, for direct comparison of the two time-points and inclusion of insulin-users, C-peptide and insulin (Elecsys Cobas, Roche diagnostics, Mannheim, Germany and Mercodia Iso-insulin ELISA, detecting mainly endogenous or both endogenous and exogenous insulin, respectively; registration: Group_A_, N=456, Group_B_, N=121; follow-up: Group_B_, N=194). Since C-peptide values were analyzed using different assays, a cubic spline was fitted to the values of individuals having measurements taken with both assays, allowing for the smooth estimation of the relationship between the assays. Cubic spline imputation (R 4.0.2, Stats Package, smooth.spline function) was then applied to predict the corresponding C-peptide values of other individuals based on the fitted spline, ensuring a more accurate comparison across the dataset.

HOMA2-B and HOMA2-IR were calculated with the HOMA calculator (V2.2.3, University of Oxford, Oxford, UK; insulin as pmol/l: 1 μU/ml = 6.00 pmol/l [19], C-peptide as nmol/l).

As a sensitivity analysis, we calculated the HOMA2-IR (INS_endo+exo_) -index using insulin values measured with the kit detecting both human insulin and insulin analogues. This correlated well with HOMA2-IR calculated with the routine measurements minimally detecting insulin analogues, among the non-insulin users (R=0.93, p=5.0748E-67), but not among the insulin users (R=-0.27, p=0.191).

### 2.5 Statistical analysis

Data are reported as median[IQR] or mean±SD, variables with skewed distributions were log_e_-transformed for linear regression analyzes. Comparisons of continuous variables between subgroups were analyzed using Kruskal-Wallis test, and intragroup changes with related samples Wilcoxon signed rank -test. Analysis of covariance (ANCOVA) was used for adjusted intergroup comparisons, and Spearmańs rank correlation coefficient for correlations. Multivariable linear regression was conducted using either stepwise forward or enter method. We report Bonferroni-corrected p-values for Kruskal-Wallis and nominal p-values for Wilcoxon signed rank -test and linear regression models (<0.05 considered statistically significant). IBM Statistics SPSS 27.0 was used for statistical analysis.

### 2.6 Data and Resource Availability

For issues of patient confidentiality and restrictions in institutional review board permissions, original de-identified data is only available through specific reasonable request to the corresponding author and material transfer agreement following EU regulations.

## 3. RESULTS

### 3.1 Changes in the cluster variables during follow-up and cluster stability (Group_A_& Group_B_)

We compared data collected at registration and at follow-up 4.6[2.8] years later for 547 individuals (40 SAID, 16 SIDD, 52 SIRD, 148 MOD, 291 MARD) (Tables S1-S6). Overall, the differences between the subgroups regarding the clustering variables diminished during the follow-up. While HbA1c was initially considerably higher in SIDD than in the other subgroups, this difference attenuated later with a major decrease in SIDD (from 11.5[1.8] to 7.4[1.7]%, 101.6[19.1] to 57.5[19.0] mmol/mol, p<0.001), and an increase in MARD (from 6.0[0.7] to 6.2[0.8]%, 42.0[8.0] to 44.0[9.0] mmol/mol, p<0.001) (Table S4, Figure 1). Also, the HOMA2-B reflecting fasting insulin secretion was lowest at registration in the SIDD group but increased during follow-up (from 36.6[25.2] to 60.2[39.9], p=0.044).

**Figure 1.**
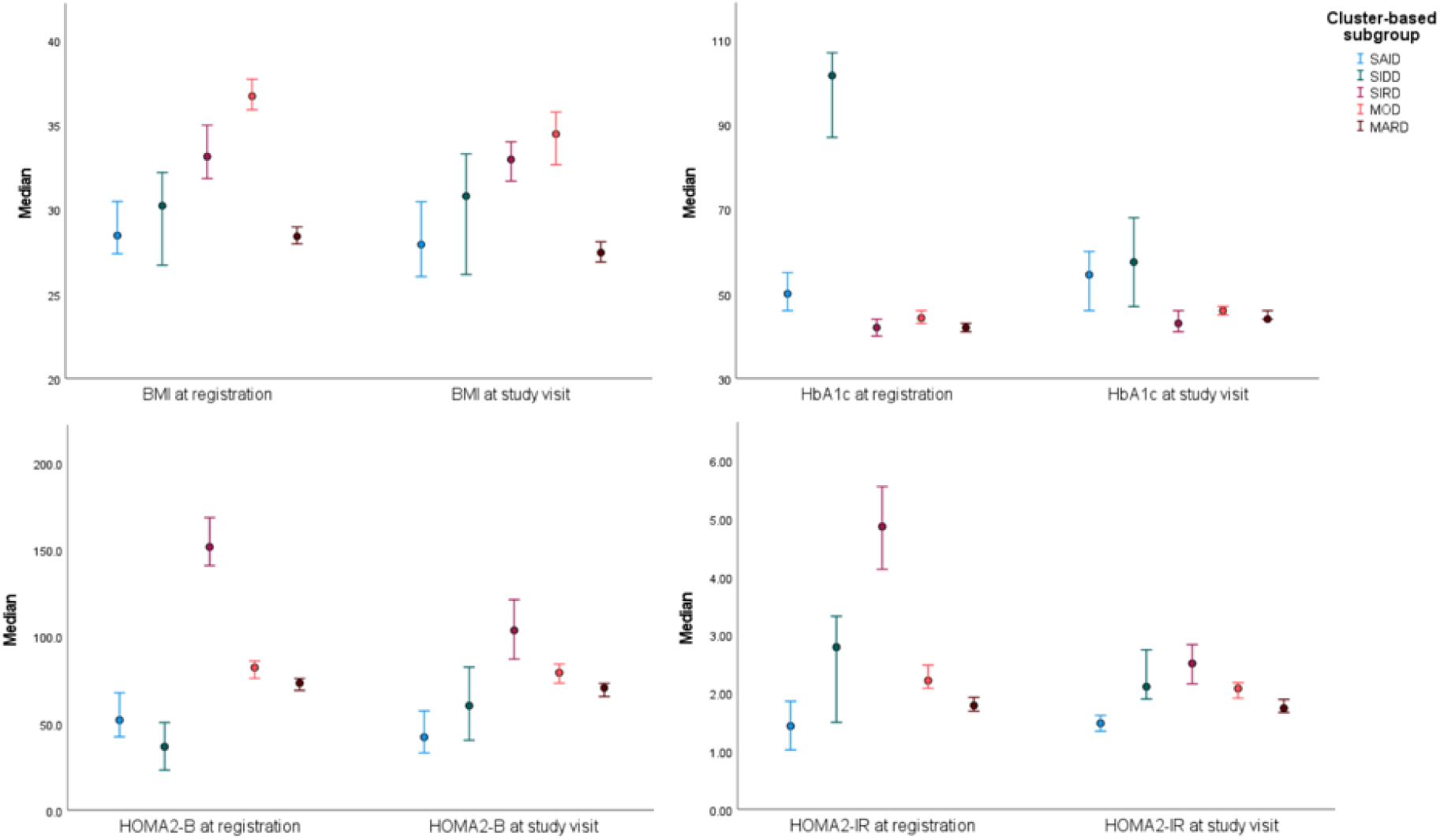
Distribution of BMI (upper left panel), HbA1c (upper right panel), HOMA2-B (lower left panel) and HOMA2-IR (lower right panel) at registration and at the study visit, stratified by the cluster-based subgroup of diabetes (see Table S4, n=547; 40 SAID, 16 SIDD, 52 SIRD, 148 MOD, 291 MARD). Data are median (95% CI). SAID/SIDD/SIRD, Severe autoimmune/insulin-deficient/insulin-resistant diabetes; MOD/MARD, mild obesity-/age-related diabetes; BMI, body mass index; HbA1c, hemoglobin A1c; HOMA2-B, HOMA2-IR, Homeostasis model assessment of β-cell function and insulin resistance, respectively.

**Figure 2.**
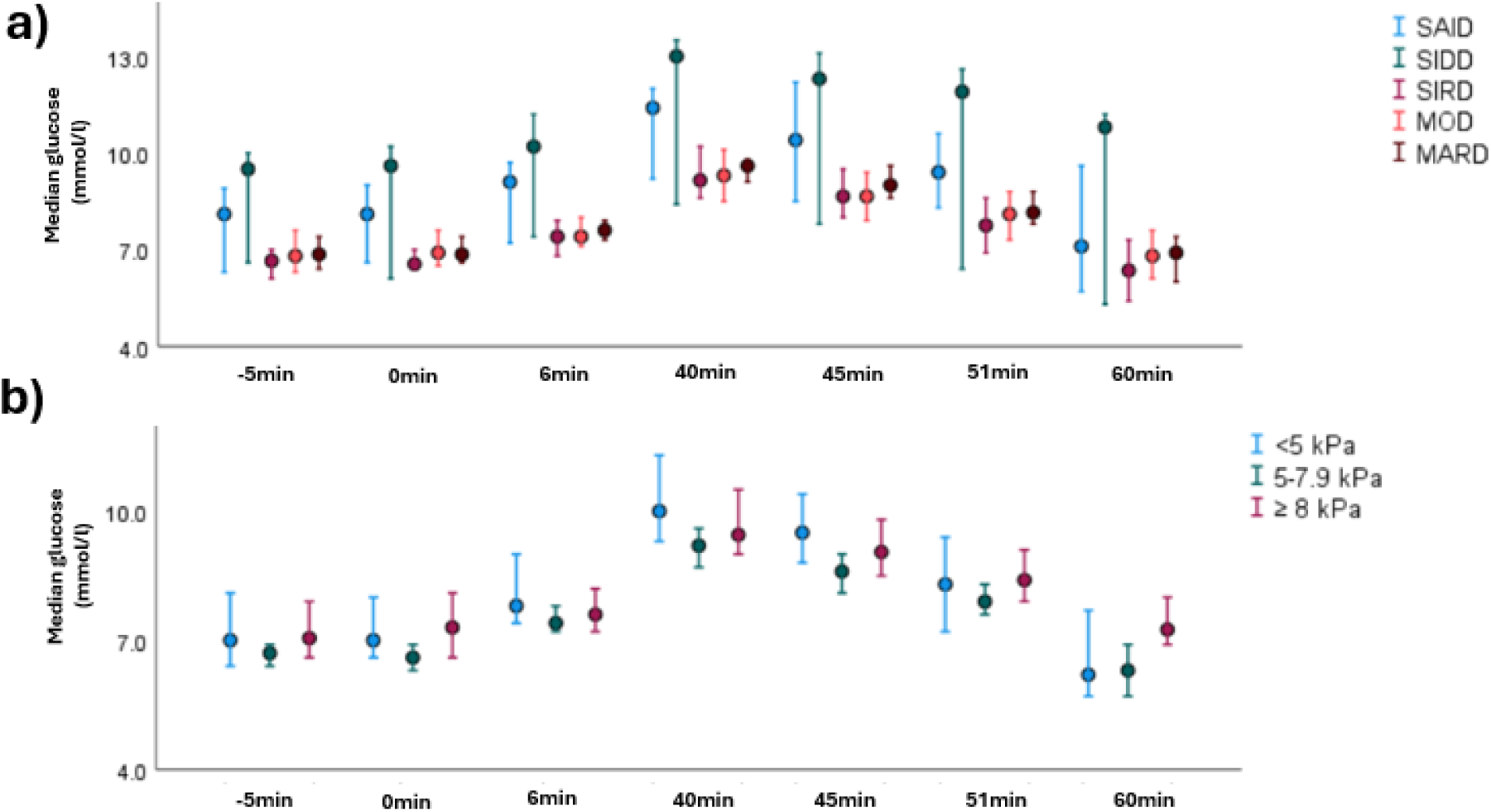
Median (95% confidence interval) plasma glucose during an i.v. glucagon-insulin tolerance test, stratified for (a) diabetes subgroup or (b) the degree of liver stiffness (LSM). 0.5 mg glucagon 0,5mg and 0.05 U/kg insulin was administered at 0 and 6 minutes, respectively. Number of patients: a) SAID n=17, SIDD n=7, SIRD n=28, MOD n=42, MARD n=60; b) LSM <5kPa n=31, LSM 5-7.9 kPa n=65, LSM ≥ 8kPa n=50. Abbreviations: SAID/SIDD/SIRD, Severe autoimmune/insulin-deficient/insulin-resistant diabetes; MOD/MARD, mild obesity-/age-related diabetes. LSM, Liver Stiffness Measurement.

On the other hand, although the level of HOMA2-B and HOMA2-IR (reflecting insulin resistance) was highest in SIRD at both time points, it decreased significantly in between (151.8[47.2] vs. 103.7[53.9], p<0.001; 4.9[2.6] vs. 2.5[1.5], p<0.001; respectively). This happened despite a stable BMI (individual level correlation between decrease in BMI and HOMA2-IR, R=0.22, p=0.117), while the minor decrease in HOMA2-IR seen in MOD (from 2.2[1.2] to 2.1[1.1], p=0.027) correlated with a decrease in BMI (R=0.39, p<0.001). Data for BMI is shown in Table S2 and S4.

We compared the cluster assignment performed on data at registration and after follow-up (Figure S3, Table S7-S8; SAID excluded, being based on GADA positivity at baseline). While most participants in the MOD (76%) and MARD (91%) groups kept their original cluster, only 13% (N=2/16) in the SIDD and 21% (N=11/52) in the SIRD group did, mostly reflecting the changes in insulin secretion and resistance indices described above (Table S8). The reallocated participants mainly moved to the MARD group [8 (50%) of SIDD, 26 (50%) of SIRD, 29 (20%) of MOD].

Compared with those remaining in the original group, those reallocated from SIRD to MOD were younger, and those from MOD to MARD older and lost more weight (Table S7).

### 3.2 Different measures of insulin secretion and sensitivity (study visit, Group_B_)

The fasting measures of insulin secretion (Table 1) correlated moderately with glucagon-stimulated increase in C-peptide (ΔC-peptide 0-6 min) (fS-C-peptide: R=0.44, p<0.001; HOMA2-B: R=0.50, p<0.001). ΔC-peptide was lowest in SAID (0.39[0.51] nmol/l, p<0.02 vs. MARD and SIRD) and SIDD (0.40[0.46] nmol/l) and highest in SIRD (0.98[0.66] nmol/l; p<0.02 vs. SAID, SIDD and MOD) (Table 1, Figure S4).

**Table 1.** Clinical characteristics of the patients participating in the study visit (Group_B_) 5.9 (95% CI 5.6-6.3) years after diagnosis, stratified by the cluster-based subgroup at registration.

| n=194 | SAID<br>(n=29) | SIDD<br>(n=11) | SIRD<br>(n=35) | MOD<br>(n=51) | MARD<br>(n=68) |
| --- | --- | --- | --- | --- | --- |
| Age years | 61.3 (25.3) | 66.5 (15.8) | 68.3 (10.6) | 59.2 (18.6) | 71.4 (8.3) |
| Duration years | 6.8 (2.5) | 5.9 (4.3) | 6.3 (2.8) | 5.9 (3.5) | 5.3 (2.2) |
| BMI kg/m<sup>2</sup> | 27.8 (6.5) | 30.2 (4.1) | 33.3 (5.2) | 34.9 (6.6) | 27.6 (4.8) |
| Female % (n) | 73 (21) | 27 (3) | 51 (18) | 57 (29) | 44 (30) |
| HbA1c % | 7.2 (2.1) | 7.5 (1.9) | 6.1 (0.8) | 6.5 (0.7) | 6.3 (0.9) |
| HbA1c mmol/mol | 55.0 (23.0) | 59.0 (21.0) | 43.0 (9.0) | 47.0 (8.0) | 45.5 (10.0) |
| Fasting glucose<br>mmol/l | 8.5 (4.1) | 9.2 (3.9) | 6.6 (1.5) | 6.9 (2.1) | 6.9 (1.7) |
| Fasting C-peptide<br>nmol/l | 0.61 (0.73) | 0.95 (0.44) | 1.47 (0.83) | 1.20 (0.55) | 1.08 (0.62) |
| Fasting Insulin μU/ml | 4.4 (9.3) | 12.4 (10.4) | 22.7 (12.2) | 15.6 (12.0) | 13.7 (12.2) |
| HOMA2-B | 41.1 (60.9) | 55.4 (23.7) | 134.4 (78.9) | 96.3 (51.5) | 90.4 (48.6) |
| HOMA2-IR | 1.6 (1.6) | 2.5 (1.3) | 3.5 (2.2) | 2.9 (1.6) | 2.7 (1.6) |
| FFA mmol/l | 642 (357) | 530 (360) | 676 (218) | 705 (235) | 649 (243) |
| Adipo-IR<br>mmol/lxμU/ml | 4.1 (5.8) | 7.3 (6.2) | 15.5 (10.9) | 10.0 (13.3) | 7.7 (10.2) |
| C-peptide response<br>nmol/l* | 0.39 (0.51) | 0.40 (0.46) | 0.98 (0.66) | 0.50 (0.54) | 0.72 (0.39) |
| C-peptide 6 min<br>nmol/l † | 1.06 (1.01) | 1.54 (0.56) | 2.51 (1.00) | 1.81 (0.81) | 1.88 (0.82) |
| K<sub>ITT</sub> %/min | 1.86 (1.33) | 1.08 (0.57) | 1.64 (1.02) | 1.68 (1.01) | 1.64 (1.04) |
| CAP dB/m | 278 (94) | 303 (78) | 321 (75) | 313 (75) | 284 (73) |
| LSM kPa | 5.5 (2.2) | 6.9 (3.6) | 7.4 (5.3) | 7.0 (4.6) | 6.2 (3.1) |
| LSM $\geq 8$kPa, % (n) | 17 (4) | 36 (4) | 42 (13) | 41 (19) | 27 (17) |
| Glucagon ng/l | 12.7 (16.2) | 22.5 (17.9) | 21.9 (14.6) | 20.8 (16.0) | 18.9 (18.4) |
| eGFR ml/min | 91 (29) | 97 (43) | 76 (36) | 96 (21) | 85 (15) |
| On insulin % (n) | 55 (16) | 55 (6) | 3 (1) | 6 (3) | 6 (4) |
| Metformin % (n) | 59 (17) | 64 (7) | 60 (21) | 82 (42) | 69 (47) |
| DPP-4i % (n) | 14 (4) | 36 (4) | 14 (5) | 16 (8) | 15 (10) |
| SGLT-2i % (n) | 14 (4) | 55 (6) | 34 (12) | 35 (18) | 24 (16) |
| GLP-1RA % (n) | 3 (1) | 46 (5) | 9 (3) | 28 (14) | 7 (5) |
| Dyslipidemia medication % (n) ‡ | 31 (9) | 64 (7) | 71 (25) | 61 (31) | 68 (46) |

The study visit occurred 5.2 (95% CI 4.8-5.8) years after registration. Data are median (IQR). SAID/SIDD/SIRD, Severe autoimmune/insulin-deficient/insulin-resistant diabetes; MOD/MARD, mild obesity-/age-related diabetes; BMI, body mass index; HbA1c, hemoglobin A1c; HOMA2-B and HOMA2-IR, Homeostasis model assessment of β-cell function and insulin resistance (calculated using C-peptide); Adipo-IR, Adipose tissue insulin resistance -index; FFA, Free fatty acid; $K_{ITT}$ , first-order rate constant for glucose disappearance; CAP, continuous attenuation parameter; LSM, Liver stiffness measurement; eGFR, estimated glomerular filtration rate; DPP-4, Dipeptidyl Peptidase 4 inhibitor; SGLT-2i, Sodium-Glucose Transport Protein 2 inhibitors; GLP-1RA, Glucagon-Like Peptide-1 Receptor Agonists. \* Change in C-peptide level 6 min after 0.5mg glucagon iv, † C-peptide 6 min after 0.5mg glucagon iv, absolute value, ‡ statin or ezetimibe.

The measures for insulin sensitivity, HOMA2-IR and the glucose disappearance rate (K_ITT_) (Table 1, Figure S4), correlated weakly (R=-0.33, p<0.001). We expected the groups with lowest HOMA2-IR to have the highest K_ITT_, but that held only for the SAID group (1.86[1.33] %/min) while the SIDD group had the lowest K_ITT_ level (1.08[0.57] %/min) (Table 1, Figure 1, Figure S4). Likewise, after adjusting with FPG and HbA1c, only SAID had better insulin sensitivity by K_ITT_ than SIRD, MOD and MARD (p<0.03, Figure S5). Of note, despite having a significantly higher HOMA2-IR, the SIRD group did not significantly differ from the other groups (except SAID) with respect to K_ITT_.

Adipose-tissue insulin resistance (Adipo-IR) was highest in SIRD and lowest in SAID (15.5[10.9] and 4.1[5.8] mmol/l x μU/ml; SIRD vs. SAID/MARD, p<0.03, SAID vs. MOD/ MARD, p<0.002; unchanged after excluding insulin users) (Table 1). At registration, the FFA level was lower in SIRD compared to SAID, MOD and MARD (319[306] vs. 542[212], 602[342], 525[303] mmol/l, p<0.01), but not after the follow-up. Also, differing from the other subgroups, FFA and HOMA2-B correlated negatively in SIRD (R=-0.41, p=0.010).

### 3.3 Liver fat content and fibrosis in the subgroups (study visit, Group_B_)

The liver fat content (CAP) was higher in both SIRD (321[75] dB/m) and MOD (313[75] dB/m) compared with SAID (278[94] dB/m; p<0.005) and MARD (284[73] dB/m; p<0.01) groups (Table 1). These differences disappeared after adjusting for BMI and glucose lowering medications (data not shown). The distribution of the LSM fibrosis measurement followed that of CAP, but with more variance and no significant differences between the groups after adjusting with BMI or glucose lowering medications (unadjusted: SIRD vs SAID; p=0.048372). Nominally, median LSM was highest (7.4[5.3] kPa) and liver fibrosis (LSM>8kPa) most prevalent (42%) in SIRD (Table 1, Figure S6).

We analyzed the BMI-independent association of measures of insulin resistance with CAP and LSM using linear regression (data not shown) within the subgroups. In SIRD, CAP was associated with Adipo-IR (β=0.564, p<0.001) and K_ITT_ (β=-0.500, p=0.002) but not with HOMA2-IR. LSM was associated with Adipo-IR and HOMA2-IR in both SIRD (β=0.651, p<0.001; β=0.424, p=0.029, respectively) and MARD (β=0.320, p=0.015; β=0.304, p=0.020, respectively), but with K_ITT_ only in MARD (β=-0.474, p<0.001).

### 3.4 Association of liver fibrosis with hepatic, adipose tissue and muscle insulin resistance (study visit, combined Group_B_)

In a multivariable logistic regression model the presence of liver fibrosis (presence/absence: LSM ≥8/<5 kPa, excluding measures with 5-8 kPa[17]) was associated with all measures of insulin sensitivity after adjustment with BMI or CAP: HOMA2-IR (OR[95% CI] 3.90[2.05-7.42; p<0.001]), Adipo-IR (1.14[1.06-1.022]; p<0.001) and K_ITT_ (0.33[0.17-0.64]; p<0.001) (Table 2).

**Table 2.** Factors associated with liver fibrosis at the study visit (N=92), according to elastography (LSM≥ 8kPa, n=57 vs. LSM < 5kPa, n=35) in a logistic regression analysis without adjustment (left), and adjusted for BMI (middle) or CAP (right).

|  |  |  |  |  | Adjustment |  |  |  |  |
| --- | --- | --- | --- | --- | --- | --- | --- | --- | --- |
|  |  | No |  |  | BMI |  |  | CAP |  |
| Variable | OR | p | AUC | OR | p | AUC | OR | p | AUC |
| Age (years) | 1.02 (0.98-1.06) | 0.371 | 0.53 (0.41-0.66) | 1.04 (0.99-1.08) | 0.119 | 0.73 (0.63-0.84) | 1.05 (1.00-1.0) | 0.052468 | 0.77 (0.67-0.87) |
| Sex (men) | 1.15 (0.49-2.67) | 0.751 | 0.52 (0.40-0.64) | 1.27 (0.51-3.15) | 0.612 | 0.73 (0.62-0.84) | 0.94 (0.37-2.41) | 0.938 | 0.75 (0.65-0.85) |
| BMI (kg/m<sup>2</sup>) | 1.16 (1.06-1.27) | 0.000844 | 0.73 (0.62-0.83) | - | - | - | 1.10 (0.99-1.21) | 0.077456 | 0.77 (0.67-0.87) |
| K<sub>ITT</sub> (%/min) | 0.33 (0.17-0.64) | 0.000918 | 0.76 (0.65-0.88) | 0.37 * (0.18-0.74) | 0.004918 * | 0.79 (0.68-0.90) | 0.38 (0.19-0.74) | 0.004644 | 0.80 (0.70-0.90) |
| HOMA2-IR | 3.90 (2.05-7.42) | 0.000035 | 0.82 (0.73-0.91) | 3.25 (1.63-6.49) | 0.000850 | 0.83 (0.75-0.92) | 2.96 (1.55-5.63) | 0.000967 | 0.84 (0.76-0.93) |
| Adipo-IR (mmol/l x μU/ml) | 1.14 (1.06-1.22) | 0.000444 | 0.75 (0.65-0.85) | 1.11 (1.03-1.20) | 0.004874 | 0.78 (0.69-0.88) | 1.10 (1.02-1.19) | 0.011857 | 0.80 (0.71-0.89) |
| fS-Insulin<sub>endo</sub> (μU/ml) | 1.12 (1.05-1.18) | 0.000326 | 0.76 (0.67-0.86) | 1.10 (1.03-1.17) | 0.003379 | 0.79 (0.69-0.88) | 1.09 (1.02-1.16) | 0.007187 | 0.81 (0.72-0.90) |
| FFA (mmol/dl) | 1.002 (1.000-1.004) | 0.058053 | 0.62 (0.50-0.73) | 1.001 (0.999-1.003) | 0.357678 | 0.74 (0.63-0.84) | 1.001 (0.999-1.003) | 0.509847 | 0.75 (0.65-0.86) |
| Triglycerides (mmol/l) | 2.28 (1.13-4.61) | 0.022187 | 0.63 (0.51-0.74) | 1.71 (0.83-3.53) | 0.146690 | 0.75 (0.64-0.85) | 1.61 (0.75-3.46) | 0.224880 | 0.76 (0.66-0.86) |
| Glucagon (ng/l) | 1.02 (0.99-1.05) | 0.154305 | 0.63 (0.51-0.75) | 1.02 (0.99-1.05) | 0.143317 | 0.74 (0.63-0.84) | 1.01 (0.98-1.04) | 0.605849 | 0.74 (0.64-0.85) |
| HbA1c (mmol/mol) | 1.02 (0.98-1.07) | 0.302829 | 0.53 (0.41-0.65) | 1.01 (0.97-1.06) | 0.611948 | 0.73 (0.62-0.83) | 1.02 (0.97-1.07) | 0.509222 | 0.76 (0.66-0.86) |
| CAP (dB/m) | 1.02 (1.01-1.03) | 0.000177 | 0.74 (0.64-0.85) | 1.01 (1.00-1.02) | 0.010803 | 0.77 (0.67-0.87) | - | - | - |

Data are shown for OR or AUC with 95% CI. Abbreviations: OR, Odds ratio; LSM, Liver stiffness measurement; SAID/SIDD/SIRD, Severe autoimmune/insulin-deficient/insulin-resistant diabetes; MOD/MARD, mild obesity-/age-related diabetes; BMI, body mass index; $K_{ITT}$ , first-order rate constant for glucose disappearance; ; HOMA2-IR, Homeostasis model assessment of insulin resistance; insulin concentration was analyzed with two methods, one mainly detecting endogenous insulin ( $INS_{endo}$ ), the other detecting both endogenous insulin and insulin analogues ( $INS_{endo+exo}$ ); FFA, Free fatty acid; HbA1c, hemoglobin A1c; CAP, continuous attenuation parameter. \* Note that the i.v. insulin was administered according to the weight (0.05 IU/kg) (see Methods).

This was consistent with analysis of covariance: mean[95% CI] K_ITT_ was significantly lower in LSM group ≥8 kPa (1.44[1.25-1.64] %/min) compared to <5 kPa (2.11[1.80-2.42] %/min; p=0.002) group even after adjusting for BMI and other confounding factors. Results using HOMA2-IR and Adipo-IR were similar to K_ITT_ (Figure 3) after excluding insulin users. Also, area under the Receiver Operating Curve increased statistically significantly when HOMA2-IR was added to a model with BMI or CAP in detection of liver fibrosis (AUC[95% CI] 0.73[0.63-0.84] vs. 0.84[0.75-0.92]; 0.75[0.65-0.85] vs. 0.85[0.77-0.93]; respectively, p<0.03 in both, Table 2, Figure S7). On the contrary, at the time of registration, only BMI and SIRD-group, but not HOMA2-IR and Adipo-IR, associated with later presence of liver fibrosis in a univariate logistic regression analysis (Tables S9).

**Figure 3.**
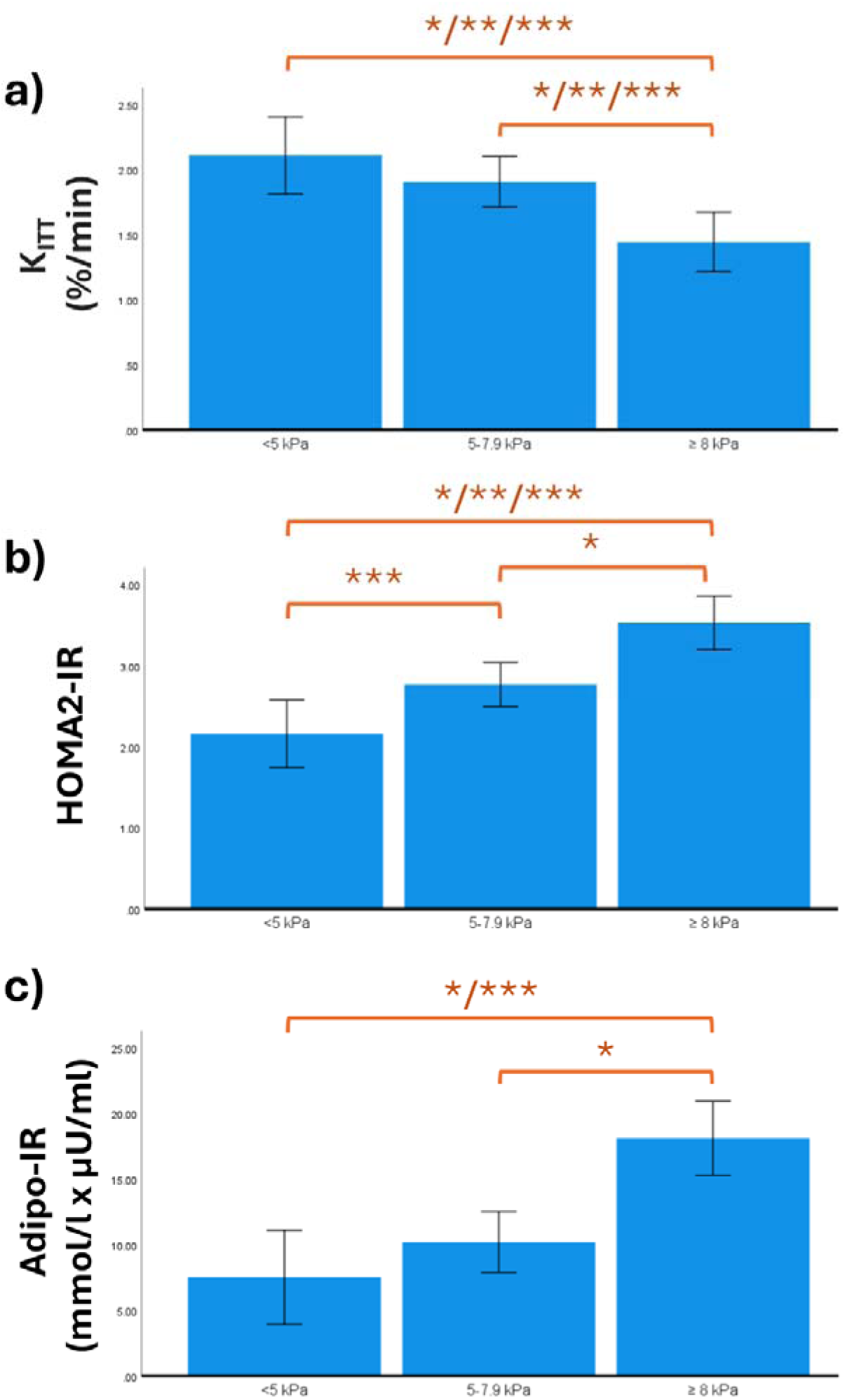
Surrogate measures of insulin resistance on the x-axis (a) K_ITT_, b) HOMA2-IR and c) Adipo-IR stratified for the severity of liver fibrosis at the study visit on the y-axis determined by liver stiffness measurement (< 5kPa, n=35; 5-7.9 kPa, n=84; ≥ 8 kPa, n=57). Abbreviations: K_ITT,_ first-order rate constant for glucose disappearance; HOMA2-IR, Homeostasis model assessment of insulin resistance; Adipo-IR, Adipose tissue insulin resistance -index. *p<0.05 with no adjustments for confounding factors, ** p<0.05 adjusted for age, sex, BMI and glucose lowering medications, *** p<0.05 adjusted for age sex, BMI and glucose lowering medications after excluding users of exogenous insulin. Analysis of covariance.

The supplement shows also linear (univariate) regression models comparing the effects of the variables for LSM and CAP (Supplement S10-S11). In brief, regarding LSM, Adipo-IR had the strongest association at the study visit and registration in the whole group, while HOMA2-IR had the strongest association when insulin users were excluded. Also fasting insulin at the study visit, detached from the theindices, showed independent association with LSM (β=0.276, p<0.001). On the contrary, BMI had the strongest association with CAP both at the registration (β=0.334, p<0.001) and at the study visit (insulin users included: β=0.334, p<0.001; insulin users excluded β=0.346, p<0.001). Multivariable regression models yielded similar results (data not shown).

## 4. CONCLUSIONS

In this regionally representative prospective study on cluster-based subgroups of adult-onset diabetes, we studied the progression of insulin deficiency and sensitivity and evaluated the stability of the subgroup assignment, as well as manifestation of insulin resistance in different tissues, focusing on liver steatosis and fibrosis. We found that, first, the overall differences in the clustering variables between subgroups diminished during the follow-up. Second, membership in the two larger subgroups (MOD, MARD) was relatively stable over time (with 76-91% remaining in the same cluster), whereas that held true for only 13-21% of the smaller subgroups (SIDD, SIRD) after a median follow-up of 4.6 years. Third, differences between the subgroups in HOMA2-indices based on fasting measures were not mirrored in the stimulated measures of insulin secretion and sensitivity, particularly regarding insulin resistance. Fourth, the overall prevalence of LF among individuals with diabetes was associated with insulin resistance in liver, muscle and adipose tissue independently of BMI or SLD. However, subgroup-specific differences in SLD and LF were mainly explained by BMI.

The changes in cluster assignment were likely affected by two observations: differences between subgroups regarding the cluster variables diminished during the follow-up, and the levels of the variables changed differently among the subgroups (Table S4). In the insulin-deficient SIDD group we observed a marked decrease in HbA1c accompanied by an increase in HOMA2-B, suggesting that the initial marked hyperglycemia and diminished insulin secretion partially reverted with commencement of treatment. In all other groups HbA1c increased during the follow-up. Of note, HOMA2-indices decreased clearly in SIRD despite stable BMI but not in MOD despite a decrease in BMI. The more frequent use of GLP-1-receptor agonists (GLP1-RA) in MOD (28% vs. 9% in SIRD) could affect the change in BMI. Altogether, glucose levels in SIRD seemed to remain lower with “lighter” treatment compared to MOD.

Our data highlights the attenuation of distinctive features of the subgroups with time and treatment. This emphasizes the importance of performing clustering in the early years after diagnosis, considering the different risk trajectories for comorbidities demonstrated previously. [2] While the results support previous data [3] on the stability of the bigger MOD and MARD clusters over time, most participants belonging to the SIDD and SIRD subgroups were reassigned to other clusters. Most were regrouped as MARD, increasing its proportion by 13% compared to registration phase. This is a logical consequence of treatment, as individuals in MARD bear the closest resemblance to those without diabetes. Also, most group changers were at the cluster borders at registration sharing some characteristics of the ‘target’ subgroup (Table S7).

The HOMA-indices are increasingly used to estimate insulin sensitivity and secretion. However, having originally been modelled in individuals without diabetes, their performance is affected by hyperglycemia, low C-peptide values [20] and use of exogenous insulin. [21] We compared the HOMA-indices with results from a combined glucagon–insulin tolerance test (GITT), previously validated against an intravenous glucose tolerance test (IVGTT) and hyperinsulinemic euglycemic clamp (HEC). [16] The glucagon-induced C-peptide secretion correlated moderately to HOMA2-B and resulted in comparable differences between the subgroups, similarly to a previous report on IVGTT. [3] In contrast, the HOMA2-IR correlated only weakly with the glucose disappearance rate K_ITT_, which is understandable as they reflect insulin resistance of the liver [22, 23], and skeletal muscle differently. Thus, differing from a previous study [3], our subgroups mainly differed regarding the liver insulin resistance but shared quite similar levels of muscle insulin sensitivity (except for SAID). Besides the different methods to evaluate insulin sensitivity, the SIRD patients in the referred HEC cohort [3] had notably higher BMI and HbA1c with younger participants and male overrepresentation.

Interestingly, at registration, median FFA level was significantly lower in SIRD than in MOD, but due to the higher fasting insulin in SIRD, the product of FFA x Insulin (Adipo-IR) was approximately similar. We speculate that the considerably lower FFA level in SIRD could indicate a reasonably effective insulin action in the prevention of lipolysis. [24] High FFA combined with low fasting insulin suggests dysfunctioning adipose tissue as the primary defect in MOD, [25] while low FFA combined with high fasting insulin pairs with insulin resistance outside of the adipose tissue in SIRD. This hypothesis is strengthened by the fact that in SIRD, unlike MOD, fasting insulin secretion (HOMA2-B) correlated with fasting FFA. Another possible explanation could be metabolic flexibility, or bodýs ability to switch between using glucose or FFA as an energy source.[26]

The liver fat content was highest in SIRD and MOD, mostly explained by BMI. The higher prevalence of SLD in these groups is in line with earlier reports mainly based on recorded ICD-codes or ALT-values [1, 2] but also MRI-data. [3] A previous retrospective study also reported that risk of liver fibrosis was associated with SIRD (although evaluation was restricted to those with prior suspicion of fibrosis). [4] In our prospective study, SIRD had highest prevalence and degree of liver fibrosis with a statistically significant difference compared with SAID. Further studies with larger unselected groups are needed to establish or refute the possible association.

Liver fibrosis was associated, independently of weight or SLD, with all measures of insulin resistance: HOMA2-IR and Adipo-IR -indices based on fasting C-peptide/insulin as well as insulin sensitivity derived from insulin tolerance test (Table 2). Interestingly, of the measurements at the time of registration, only Adipo-IR and BMI were associated with later liver fibrosis (Table S11), seemingly preceding liver fibrosis. This is in line with a previous study suggesting Adipo-IR to predict the severity of liver fibrosis in individuals with type 2 diabetes and SLD. [27] Taken together, our findings suggest that although BMI and SLD are known contributors to insulin resistance and liver fibrosis, [10, 28] they do not comprehensively explain the association between insulin resistance in different organs and liver fibrosis in individuals with diabetes. Of note, the degree of steatohepatitis cannot be measured without a biopsy.

Both HOMA2-IR and Adipo-IR -indices incorporate fasting insulin, and hyperinsulinemia has been implicated in liver fibrosis via activation of hepatic stellate cells, increased collagen-production and accumulation of extracellular matrix. [29, 30] In our study, also fasting insulin showed association with liver fibrosis, when analyzed separately from the indices (Table S11). However, we cannot discern whether high insulin concentration only reflects the overall insulin resistance or has an independent role in disease progression.

Strengths of this study include regional coverage of one of the original cohorts reporting the subtypes [1], and the prospective study setting. Using VCTE instead of liver biopsy can be considered both a strength and a limitation. VCTE estimates a larger volume of liver than biopsy, and pathological interpretation of liver biopsies can be subjective. Also, VCTE is widely used in clinical practice while only a minority of individuals with SLD ever undergo liver biopsy. On the other hand, despite comprehensive validation studies [17], VCTE may not give the same level of accuracy as liver biopsy, it does not measure steatohepatitis, and a high CAP value may result in exaggerated level of LSM. [31] However, in our study, adjusting LSM with CAP did not affect the results. Limitations also include the relatively small dataset and vulnerability to statistical type II errors, especially in the SIDD group (n=16). Results of this group should be interpreted with caution. Short follow-up time could be also considered a limitation since the development of liver fibrosis can take decades. [32] Also, we did not follow the use of medications between registration and study visit. We also want to point out that the subgroups defined by hard clustering cannot be directly implemented in the clinic or for individual cases. Instead, their usefulness is in studying patterns and creating hypotheses.

In conclusion, we showed that although the differences became less distinctive, the subgroups of diabetes still differed in insulin secretion and resistance after few years of disease burden. We speculate that the odds of being assigned to the same cluster probably weaken with longer follow-up, perhaps due to the natural heterogeneity in disease progression and different treatments. The increasing interventional possibilities highlight the importance of finding individuals at risk of having SLD or its progressed forms. [12, 13, 15, 33, 34] The association of insulin resistance in different tissues with the occurrence of liver fibrosis, independently from BMI or liver steatosis, as well as the reports on increased risk of comorbidities in the most insulin resistant group (SIRD) irrespective of glucose control [2] speaks for screening for insulin resistance in all individuals with type 2 diabetes.

Although the subgroups used in this study are not comprehensive, they reflect the heterogeneity of the disease and provide a stepping stone for future studies. Controlled randomized trials of different medication strategies as well as prospective studies with longer follow-up would provide more information about subgroups and assistance for clinical decisions in future.

## 5. ARTICLE INFORMATION

## 5.1 Acknowledgements

We gratefully acknowledge the contribution of the founder of the DIREVA Study, professor Leif Groop, and the study nurses and research assistants Matleena Lamminaho, Linda Sjölund, Britt Stolpe, and Paula Kokko, as well as the DIREVA and Botnia study groups.

## 5.3 Author contributions

A.L. and A.E.K. were involved in study design, data collection, data interpretation and manuscript composition, A.K. was involved in data collection and reviewing the manuscript, L.H. in study design and reviewing the manuscript, S.K. in data interpretation and editing, J.H. in data collection, K.L. in study design and data interpretation, M.L. and E.A. in data interpretation, editing and reviewing the manuscript. T.T. is the guarantor of this work and, as such, had full access to all the data in the study and takes responsibility for the integrity of the data and the accuracy of the data analysis.

## 5.4 Prior Presentation

Part of the results has been presented at the annual meeting of the European Association for the Study of Diabetes in 2023. A non–peer-reviewed version of this article was submitted to the MedRxiv preprint server (https://www.medrxiv.org/content/10.64898/2025.12.01.25340818v1) on 1st December 2025.

