## Supplementary data for "Subgroups of adult-onset diabetes: a prospective follow-up study of progression of insulin resistance and deficiency and association with liver steatosis and fibrosis"

**Supplement**

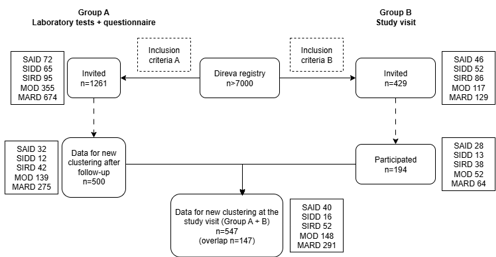

**Figure S1a. Flowchart of the follow-up study design**. Inclusion criteria for Groups A and B: 1) age at diagnosis of diabetes ≥18 years, 2) registration in DIREVA within 3 years from the diagnosis and 2-10 years (A) or 3-10 years (B) before the study, 3) available data for all variables needed for assigning the cluster subtypes. In addition, criteria for B included age under 80 years at the time of study. SAID/SIDD/SIRD, Severe autoimmune/insulin-deficient/insulin-resistant diabetes; MOD/MARD, mild obesity-/age-related diabetes. See Supplementary Table S1 and S2 for clinical characteristics at registration for the invited group (N=1319) and the included group (N=547), respectively. The flow chart was made with diagrams.net (app.diagrams.net).

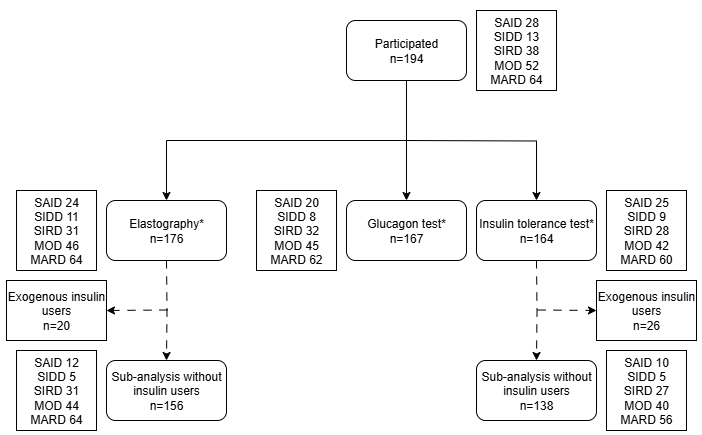

**Figure S1b. Flowchart of the follow-up study visit (Group_B_) design**. SAID/SIDD/SIRD, Severe autoimmune/insulin-deficient/insulin-resistant diabetes; MOD/MARD, mild obesity-/age-related diabetes. * = Missing data: elastography n=10, glucagon test n=27, insulin tolerance test n=19. Additionally, we excluded data for 8 individuals for elastography (7 due to prior obesity surgery, 1 due to liver metastasis), and 11 for insulin tolerance test (8 due to prior obesity surgery, 3 due to being extreme outliers: 1 SAID [K_ITT_ 0.10 %/min], 1 MOD [-0.57 %/min], 1 MARD [0.01 %/min]. The flow chart was made with diagrams.net (app.diagrams.net).

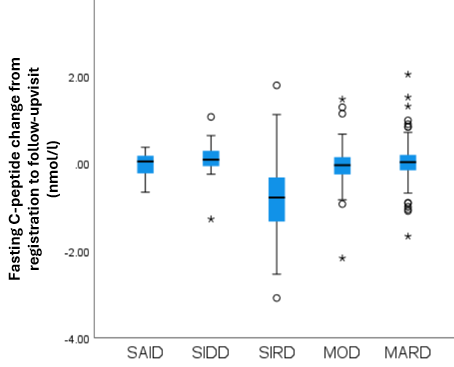

**Figure S2. The change (median and IQR) in fasting C-peptide from registration to the follow-up visit (N=547; SAID 40, SIDD 16, SIRD 52, MOD 148, MARD 291). The decrease in C-peptide was significantly higher in the SIRD group (-0.79 [IQR 1.06] nmol/l) compared to the other groups (p<0.001, Kruskal-Wallis test).** Abbreviations: SAID/SIDD/SIRD, Severe autoimmune/insulin-deficient/insulin-resistant diabetes; MOD/MARD, mild obesity-/age-related diabetes. The circles represent mild (≤3 x IQR) and asterisks extreme (>3 x IQR) outliers.

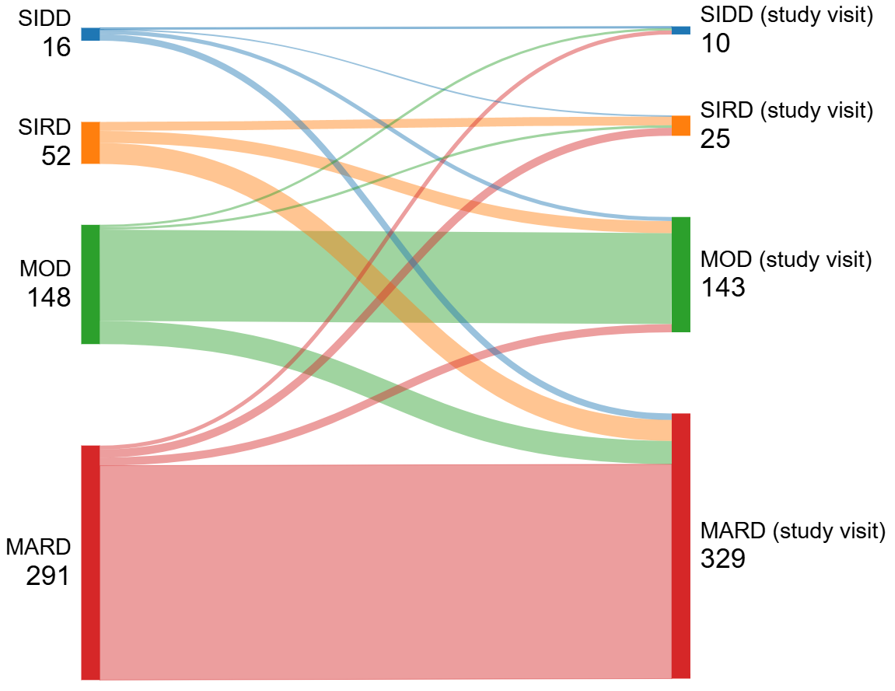

**Figure S3. Cluster distribution at registration and at the follow-up study visit.** The Sankey diagram shows the redistribution of the cluster subgroups from registration to the follow-up visit. Altogether 392/507 patients (77%) were allocated to the same cluster at registration and at follow-up (SIDD 13%, SIRD 21%, MOD 76%, MARD 91%). Assignment to SAID group was based on GADA at registration (excluded from the figure). Abbreviations: SAID/SIDD/SIRD, Severe autoimmune/insulin-deficient/insulin-resistant diabetes; MOD/MARD, mild obesity-/age-related diabetes; GADA, glutamic acid decarboxylase antibodies. Sankey diagram was made with sankeyMATIC (sankeymatic.com).

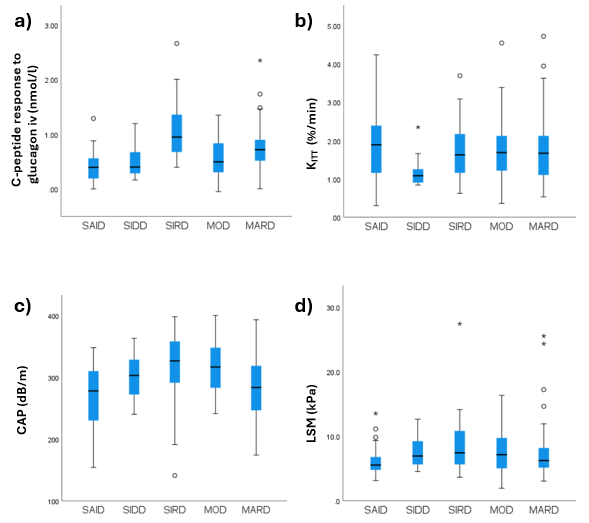

**Figure S4. Measurements of insulin secretion (a) and sensitivity (b) derived from i.v. glucagon-insulin tolerance test together with continuous attenuation parameter (CAP, c) and liver stiffness measurement (LSM, d) from liver elastography stratified by the subgroups of adult-onset diabetes.** a) 6-minute C-peptide response (nmol/l) to i.v. 0.5mg glucagon; b) K_ITT_ (first-order rate constant for glucose disappearance after i.v. insulin, %/min); c) CAP (continuous attenuation parameter, dB/m) and d) LSM (liver stiffness measurement, kPa) for assessment of liver steatosis and liver fibrosis, respectively. The circles represent mild (≤3 x IQR) and asterisks extreme (>3 x IQR) outliers. SAID/SIDD/SIRD, Severe autoimmune/insulin-deficient/insulin-resistant diabetes; MOD/MARD, mild obesity-/age-related diabetes.

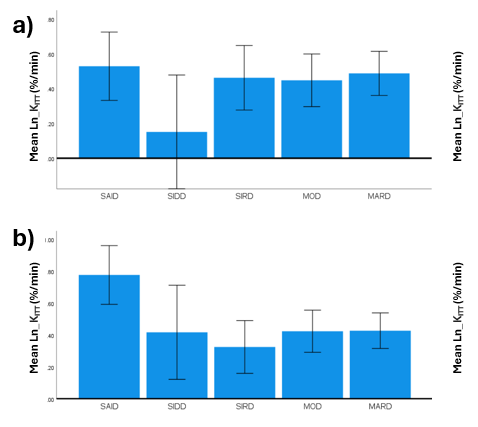

**Figure S5. The glucose disposal rate during an insulin tolerance test (K_ITT_) stratified for the cluster-based subgroups of diabetes.** The glucose-disposal rate is shown without adjustments (a) and adjusted for fasting glucose and HbA1c (b). Statistically significant differences (analysis of covariance): a) none; b) SAID vs. SIRD (P=0.006823), MOD (P=0.027625), or MARD (P=0.020111). SAID/SIDD/SIRD, Severe autoimmune/insulin-deficient/insulin-resistant diabetes; MOD/MARD, mild obesity-/age-related diabetes; K_ITT_, first-order rate constant for glucose disappearance; HbA1c, hemoglobin A1c.

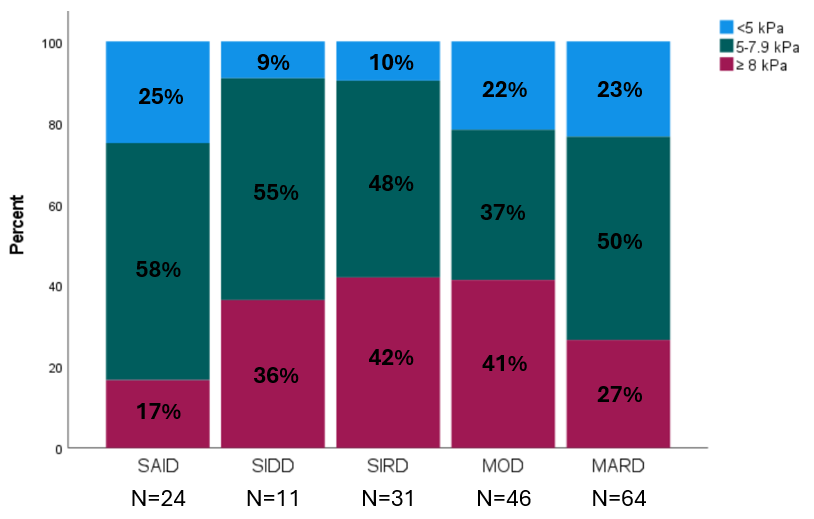

**Figure S6. Proportion of individuals with different stages of liver fibrosis by elastography (LSM<5 kPa, in blue; 5-7.9 kPa, in green; >8 kPa, in burgundy) according to the subgroup of diabetes.** No statistically significant differences using Fisher´s exact test (p=0.284775). Abbreviations: LSM, Liver stiffness measurement; SAID/SIDD/SIRD, Severe autoimmune/insulin-deficient/insulin-resistant diabetes; MOD/MARD, mild obesity-/age-related diabetes.

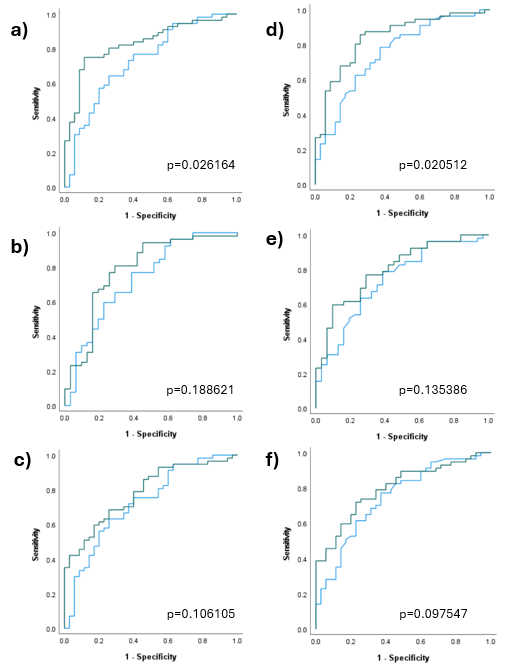

**Figure S7. Detection of liver fibrosis (Liver stiffness measurement ≥8 kPa [n=57] vs. <5kPa [n=35] in elastography: Comparison of the area under the Receiver Operating Characteristics (ROC) curves for body-mass index (BMI, panels a-c, blue lines) or liver steatosis (CAP, panels d-f, blue lines) alone vs. in models with measures of insulin resistance/sensitivity (green lines; a) BMI + HOMA2-IR, b) BMI + K_ITT_ , c) BMI + Adipo-IR, d) CAP + HOMA2-IR, e) CAP + K_ITT,_  f) CAP and Adipo-IR.** a) AUC (95% CI) 0.73 (0.63-0.84) vs. 0.84 (0.75-0.92); b) 0.75 (0.64-0.85) vs. 0.80 (0.70-0.90); c) 0.73 (0.63-0.83) vs. 0.79 (0.70-0.88); d) 0.75 (0.65-0.85) vs. 0.85 (0.77-0.93); e) 0.74 (0.64-0.85) vs. 0.80 (0.70-0.89); f) 0.75 (0.65-0.85) vs. 0.80 (0.71-0.89). Sensitivity is shown on the y-axis and specificity on the x-axis**.** CAP, continuous attenuation parameter; HOMA2-IR, Homeostasis model assessment of insulin resistance; K_ITT_, first-order rate constant for glucose disappearance; Adipo-IR, Adipose tissue insulin resistance -index.

**Table S1. Clinical characteristics at registration of all invited patients (n=1319) stratified for the cluster-based subgroup of diabetes**

|  | SAID (n=78) | SIDD (n=71) | SIRD (n=109) | MOD (n=366) | MARD (n=695) |
| --- | --- | --- | --- | --- | --- |
| Female % (n) | 50 (39) | 25 (18) | 45 (49) | 52 (190) | 40 (276) |
| Age at dg* years | 53.9 (16.5) | 53.3 (13.4) | 65.8 (9.4) | 49.4 (10.5) | 67.0 (8.1) |
| BMI kg/m2 | 29.2 (4.9) | 28.8 (5.2) | 34.1 (5.4) | 37.4 (6.3) | 28.9 (4.0) |
| HbA1c mmol/mol | 54.1 (16.6) | 91.9 (19.0) | 46.7 (14.6) | 48.9 (12.1) | 44.5 (7.8) |
| HbA1c % | 7.1 (1.5) | 10.6 (1.7) | 6.4 (1.3) | 6.6 (1.1) | 6.2 (0.7) |
| HOMA2-B^#^ | 52.9 (50.0) | 31.9 (24.0) | 142.0 (48.8) | 78.4 (40.1) | 71.1 (31.8) |
| HOMA2-IR^#^ | 1.4 (1.3) | 2.1 (2.1) | 4.9 (3.2) | 2.4 (1.2) | 1.8 (1.0) |
| On insulin % (n) | 28 (22) | 49 (35) | 10 (11) | 6 (22) | 6 (40) |

*diagnosis; data are mean (SD) or median^#^ (IQR). SAID/SIDD/SIRD, Severe autoimmune/insulin-deficient/insulin-resistant diabetes; MOD/MARD, mild obesity-/age-related diabetes; BMI, body mass index; HbA1c, hemoglobin A1c; HOMA2-B or HOMA2-IR, Homeostasis model assessment of β-cell function or insulin resistance. Altogether 1319 individuals (1261 to Group_A_ and 429 to Group_B_ with an overlap of 371 individuals).

**Table S2. Clinical characteristics at registration of the participants (with complete data for reclustering after follow-up, n=547) stratified for the cluster-based subgroup of diabetes**

|  | SAID  (n=40) | SIDD  (n=16) | SIRD  (n=52) | MOD (n=148) | MARD (n=291) |
| --- | --- | --- | --- | --- | --- |
| Proportion of those invited (%) | 51 | 23 | 48 | 40 | 42 |
| Female % (n) | 68 (27) | 31 (5) | 48 (25) | 61 (90) | 48 (140) |
| Age at dg* years | 56.7 (22.7) | 60.7 (21.3) | 63.1 (11.4) | 51.6 (14.1) | 66.8 (8.8) |
| BMI kg/m2 | 28.5 (5.3) | 30.2 (5.3) | 33.1 (7.0) | 36.7 (6.9) | 28.4 (5.2) |
| HbA1c mmol/mol | 50.0 (19.0) | 101.6 (19.1) | 42.0 (6.8) | 44.3 (10.0) | 42.0 (8.0) |
| HbA1c % | 6.7 (1.7) | 11.5 (1.8) | 6.0 (0.6) | 6.2 (0.9) | 6.0 (0.7) |
| HOMA2-B | 51.9 (49.0) | 36.6 (25.2) | 151.8 (47.2) | 82.2 (37.4) | 73.3 (33.4) |
| HOMA2-IR | 1.4 (1.5) | 2.8 (1.7) | 4.9 (2.6) | 2.2 (1.2) | 1.8 (1.1) |
| fS-Insulin_endo_ μU/ml ^#^ | 11.2 (9.8) | 16.4 (21.5) | 42.6 (44.1) | 22.3 (19.1) | 14.5 (12.5) |
| fS-Insulin_endo+exo_ μU/ml | 9.6 (17.4) | 15.2 (15.0) | 31.1 (31.3) | 14.8 (11.8) | 9.7 (7.8) |
| Fasting C-peptide nmol/l ^#^ | 0.57 (0.70) | 1.09 (0.98) | 2.46 (1.10) | 1.32 (0.65) | 1.08 (0.60) |
| FFA mmol/l ^#^ | 542 (212) | 647 (565) | 319 (306) | 602 (342) | 525 (303) |
| On insulin % (n) | 25 (10) | 50 (8) | 14 (7) | 3 (5) | 4 (11) |

*diagnosis; data are median (IQR). SAID/SIDD/SIRD, Severe autoimmune/insulin-deficient/insulin-resistant diabetes; MOD/MARD, mild obesity-/age-related diabetes; BMI, body mass index; HbA1c, hemoglobin A1c; HOMA2-B or HOMA2-IR, Homeostasis model assessment of β-cell function or insulin resistance; insulin concentration was analyzed with two methods, one mainly detecting endogenous insulin (INS_endo_ ), the other detecting both endogenous insulin and insulin analogues (INS_endo+exo_); FFA, Free fatty acid. ^#^ N= 429 patients (29 SAID, 8 SIDD, 38 SIRD, 113 MOD, 239 MARD).

**Table S3. Clinical characteristics at follow-up of the participants (with complete data for clustering, n=547) stratified for the cluster-based subgroup of diabetes**

|  | SAID  (n=40) | SIDD  (n=16) | SIRD  (n=52) | MOD (n=148) | MARD (n=291) |
| --- | --- | --- | --- | --- | --- |
| Female % (n) | 68 (27) | 31 (5) | 48 (25) | 61 (90) | 48 (140) |
| Age at diagnosis years | 63.6 (23.1) | 67.8 (19.0) | 68.0 (11.6) | 57.4 (13.8) | 72.0 (9.0) |
| Diabetes duration years | 6.1 (2.9) | 5.8 (4.1) | 6.1 (2.8) | 5.4 (3.1) | 4.9 (2.9) |
| BMI kg/m2 | 27.9 (6.0) | 30.8 (6.4) | 33.0 (5.1) | 34.5 (7.7) | 27.4 (4.9) |
| HbA1c mmol/mol | 54.5 (20.0) | 57.5 (19.0) | 43.0 (11.0) | 46.0 (10.0) | 44.0 (9.0) |
| HbA1c % | 7.1 (1.8) | 7.4 (1.7) | 6.1 (1.0) | 6.4 (0.9) | 6.2 (0.8) |
| HOMA2-B | 42.0 (36.1) | 60.2 (39.9) | 103.7 (53.9) | 79.3 (39.3) | 70.5 (33.2) |
| HOMA2-IR | 1.5 (0.5) | 2.1 (0.8) | 2.5 (1.5) | 2.1 (1.1) | 1.7 (1.0) |
| LDL mmol/l | 2.6 (1.1) | 2.5 (2.3) | 2.2 (1.0) | 2.3 (1.2) | 2.3 (1.2) |
| Triglycerides mmol/l | 0.95 (0.81) | 2.00 (1.64) | 1.68 (0.82) | 1.55 (0.94) | 1.36 (0.91) |
| eGFR ml/min | 93 (26) | 81 (41) | 78 (38) | 94 (18) | 80 (20) |

Data are median (IQR). Abbreviations: SAID/SIDD/SIRD, Severe autoimmune/insulin-deficient/insulin-resistant diabetes; MOD/MARD, mild obesity-/age-related diabetes; BMI, body mass index; HbA1c, hemoglobin A1c; HOMA2-B or HOMA2-IR, Homeostasis model assessment of β-cell function or insulin resistance; LDL, Low density lipoprotein; eGFR, estimated glomerular filtration rate.

**Table S4. The median (IQR) within-patient change in the clinical characteristics of the participants (n=547) from registration to the follow-up stratified for the cluster-based subgroup of diabetes (see Figure S2 for differences in group medians)**

|  | SAID  (n=40) | SIDD  (n=16) | SIRD  (n=52) | MOD (n=148) | MARD (n=291) |
| --- | --- | --- | --- | --- | --- |
| BMI kg/m2 | -0.3 (3.5) | +1.4 (2.1)  p=0.022895 | -0.3 (3.0) | -2.0 (4.0)  p=2.931E-14 | -0.8 (2.0)  p=3.9524E-14 |
| HbA1c mmol/mol | +1.4 (14.8) | -40.5 (29.0)  p=0.000805 | +1.5 (10.5) | +0.8 (9.0) | +2.0 (6.0)  p=2.0082E-12 |
| HbA1c % | +0.1 (1.4) | -3.7 (2.7) | +0.1 (1.0) | +0.1 (0.8) | +0.2 (0.6) |
| HOMA2-B | -2.2 (27.9) | +21.4 (44.7)  p=0.043733 | -48.2 (66.5)  p=0.000003 | -3.4 (35.3) | -0.7 (32.3) |
| HOMA2-IR | +0.1 (1.1) | -0.1 (1.4) | -1.9 (3.2)  p=1.4827E-7 | -0.1 (1.1)  p=0.027303 | +0.1 (0.9) |

Data are median (IQR). Abbreviations: SAID/SIDD/SIRD, Severe autoimmune/insulin-deficient/insulin-resistant diabetes; MOD/MARD, mild obesity-/age-related diabetes; BMI, body mass index; HbA1c, hemoglobin A1c; HOMA2-B or HOMA2-IR, Homeostasis model assessment of β-cell function or insulin resistance. P-values <0.05 (Wilcoxon signed rank test) of changes in group medians are reported.

**Table S5****.** **Clinical characteristics at registration of the patients (n=194) participating in the study visit (Group_B_) stratified for the cluster-based subgroup of diabetes**

|  | SAID  (n=29) | SIDD  (n=11) | SIRD  (n=35) | MOD  (n=51) | MARD (n=68) |
| --- | --- | --- | --- | --- | --- |
| Female % (n) | 73 (21) | 27 (3) | 51 (18) | 57 (29) | 44 (30) |
| Age years | 57.0 (25.6) | 57.4 (12.6) | 63.3 (10.3) | 55.5 (19.4) | 65.8 (9.3) |
| Age at dg* years | 54.3 (25.5) | 56.9 (13.3) | 63.1 (10.0) | 55.1 (19.8) | 65.3 (9.0) |
| BMI kg/m2 | 28.3 (5.7) | 30.0 (4.9) | 33.1 (6.6) | 37.1 (5.5) | 28.9 (5.7) |
| HbA1c mmol/mol | 51.0 (22.1) | 89.1 (25.4) | 42.1 (6.4) | 43.0 (10.0) | 41.5 (8.8) |
| HbA1c % | 6.8 (2.0) | 10.3 (2.3) | 6.0 (0.6) | 6.1 (0.9) | 5.9 (0.8) |
| fP-glucose mmol/l | 7.2 (3.3) | 12.5 (5.7) | 6.9 (1.6) | 7.2 (1.7) | 6.8 (1.3) |
| fS-C-pept nmol/l ^#^ | 0.65 (0.67) | 0.76 (0.50) | 2.12 (0.91) | 0.92 (0.43) | 0.73 (0.42) |
| fS-Ins_endo_ μU/ml ^#^ | 11.2 (14.2) | 16.4 (14.4) | 57.8 (43.2) | 20.1 (19.4) | 15.1 (19.4) |
| fS-Ins_endo+exo_ μU/ml ^#^ | 13.1 (19.3) | 12.4 (14.2) | 42.0 (32.4) | 13.5 (11.1) | 10.9 (6.7) |
| HOMA2-B_C-pept_ | 57.6 (49.3) | 35.5 (19.1) | 158.6 (40.7) | 82.8 (41.9) | 71.8 (34.7) |
| HOMA2-IR_C-pept_ | 1.8 (1.8) | 2.7 (1.7) | 5.2 (2.5) | 2.3 (1.0) | 1.8 (1.0) |
| HOMA2-B _INSendo_^#^ | 61.4 (59.6) | 29.7 (24.2) | 219.7 (104.2) | 95.9 (85.9) | 83.0 (54.7) |
| HOMA2-IR_INSendo_^#^ | 1.9 (3.2) | 2.9 (2.7) | 6.2 (4.7) | 2.8 (3.0) | 2.1 (1.2) |
| HOMA2-B_INSendo+exo_^#^ | 61.3 (41.1) | 19.8 (16.4) | 146.7 (79.5) | 64.7 (49.5) | 64.4 (34.3) |
| HOMA2-IR_INSendo+exo_^#^ | 1.7 (5.0) | 1.9 (2.0) | 4.4 (3.4) | 1.7 (1.5) | 1.4 (0.8) |
| FFA mmol/l ^#^ | 546 (201) | 645 (692) | 315 (293) | 670 (343) | 472 (323) |
| Adipo-IR mmol/lxμU/ml ^#^ | 6.2 (7.8) | 8.8 (16.2) | 14.8 (17.1) | 14.4 (13.9) | 7.0 (7.7) |
| Adipo-IR_INSendo+exo_^#^  mmol/lx μU/ml | 6.8 (9.2) | 9.6 (13.4) | 10.1 (12.5) | 9.9 (10.7) | 5.1 (4.8) |
| On insulin% (n) | 53 (9) | 24 (4) | 11 (4) | 0 (NA) | 0 (NA) |

*diagnosis, data are median (IQR). SAID/SIDD/SIRD, Severe autoimmune/insulin-deficient/insulin-resistant diabetes; MOD/MARD, mild obesity-/age-related diabetes; BMI, body mass index; HbA1c, hemoglobin A1c; HOMA2-B or HOMA2-IR, Homeostasis model assessment of β-cell function or insulin resistance; insulin concentration was analyzed with two methods, one mainly detecting endogenous insulin (INS_endo_ ), the other detecting both endogenous insulin and insulin analogues (INS_endo+exo_); Adipo-IR, Adipose tissue insulin resistance -index; FFA, Free fatty acids. ^#^N=119 patients (21 SAID, 6 SIDD, 23 SIRD, 28 MOD, 41 MARD).

**Table S6. Supplementary information to Table 1 on clinical characteristics at the study visit (Group_B_ n=194) stratified for the cluster-based subgroup of diabetes.**

|  | SAID  (n=29) | SIDD  (n=11) | SIRD  (n=35) | MOD  (n=51) | MARD  (n=68) |
| --- | --- | --- | --- | --- | --- |
| fS-Ins_endo+exo_ μU/ml | 11.5 (61.1) | 16.3 (22.5) | 15.3 (9.4) | 12.2 (8.6) | 9.9 (9.9) |
| HOMA2-BINS_endo_ | 28.3 (37.2) | 46.2 (40.7) | 112.6 (76.2) | 80.1 (44.9) | 75.9 (47.4) |
| HOMA2-BINS_endo+exo_ | 56.0 (92.3) | 49.6 (37.7) | 80.5 (59.5) | 59.0 (32.4) | 54.3 (32.6) |
| HOMA2-IRINS_endo_ | 0.7 (1.2) | 2.0 (1.3) | 3.2 (1.8) | 2.1 (1.8) | 1.9 (1.7) |
| HOMA2-IRINS_endo+exo_ | 1.6 (8.9) | 1.7 (1.8) | 1.8 (1.0) | 1.5 (1.0) | 1.2 (1.1) |
| FFA response mmol/l * | -31 (143) | 47 (161) | -16 (81) | -30 (87) | -1 (109) |
| FFA 6min mmol/l † | 636 (290) | 577 (233) | 681 (217) | 683 (233) | 655 (273) |
| Adipo-IRINS_endo+exo_  mmol/lx μU/ml | 8.5 (27.8) | 14.1 (30.9) | 9.1 (9.0) | 8.0 (8.4) | 5.3 (7.2) |
| CAP ≥ 288 dB/m, % (n) ‡ | 42 (11) | 73 (8) | 76 (25) | 68 (34) | 50 (32) |
| CAP ≥ 302 dB/m, % (n) ‡ | 31 (8) | 55 (6) | 70 (23) | 60 (30) | 44 (28) |

The study visit occurred 5.2 (95% CI 4.8-5.8) years after registration. Data are median (IQR). SAID/SIDD/SIRD, Severe autoimmune/insulin-deficient/insulin-resistant diabetes; MOD/MARD, mild obesity-/age-related diabetes; HOMA2-B or HOMA2-IR, Homeostasis model assessment of β-cell function or insulin resistance calculated with fasting insulin (instead of C-peptide as in Table 1) analyzed with two methods, one mainly detecting endogenous insulin (INS_endo_ ), the other detecting both endogenous insulin and insulin analogues (INS_endo+exo_); Adipo-IR, Adipose tissue insulin resistance -index; FFA, Free fatty acid; CAP, continuous attenuation parameter. * Change in FFA level 6 min after 0,5mg glucagon iv, † FFA concentration 6 min after 0.5mg glucagon iv, ‡ n=184 (26 SAID, 11 SIDD, 33 SIRD, 50 MOD, 64 MARD).

**Table S7. Data for the variables used for the cluster-based subgrouping of the participants at the time of registration stratified according to the cluster subgroup assigned at registration and at follow-up (first column).**

| Cluster subgroup at registration /follow-up (n) | Cluster variables at registration, median (IQR) | | | | |
| --- | --- | --- | --- | --- | --- |
|  | **BMI**  **(kg/m^2^)** | **Age at dg (years)** | **HbA1c (mmol/mol)**  **(%)** | **HOMA2-B** | **HOMA2-IR** |
| SIDD /SIDD (2) | **29.7/32.7†** | **58.6/56.9†** | **103.3/68.0†**  **11.6/8.4†** | **37.6/32.1†** | **2.7/2.2†** |
| /SIRD (1) | 34.1† | 71.7† | 103.0†  11.6† | 101.6† | 2.7† |
| /MOD (5) | 31.0 (2.1) | 49.4 (11.2) | 89.1 (20.0)  10.3 (1.8) | 30.2 (26.6) | 3.2 (2.2) |
| /MARD (8) | 26.9 (7.2) | 66.5 (12.7) | 103.3 (18)  11.6 (1.7) | 41.8 (30.1) | 3.0 (2.2) |
| SIRD /SIRD (11) | **35.0 (4.8)** | **63.8 (6.5)** | **42.1 (9.0)**  **6.0 (0.8)** | **168.7 (48.6)** | **4.2 (2.4)** |
| /MOD (15) | 35.4 (7.2) | 54.5 (10.7)  p=0.017512* | 42.0 (5.0)  6.0 (0.5) | 158.6 (81.1) | 5.9 (2.0) |
| /MARD (26) | 31.7 (5.0) | 67.0 (9.8) | 40.5 (7.0)  5.9 (0.6) | 147.4 (45.1) | 4.3 (2.8) |
| MOD /MOD (113) | **37.0 (6.7)** | **50.2 (13.9)** | **43.0 (8.0)**  **6.1 (0.7)** | **82.8 (38.3)** | **2.2 (1.1)** |
| /SIDD (3) | 28.3-40.4‡ | 33.3-64.1‡ | 43.2-46.0‡  6.1-6.4‡ | 48.0-132.2‡ | 1.3-2.5‡ |
| /SIRD (3) | 33.3-41.0‡ | 43.8-67.1‡ | 40.0-66.0‡  5.8-8.2‡ | 59.0-126.0‡ | 3.3-4.2‡ |
| /MARD (29) | 35.8 (5.7) | 57.3 (11.1)  p=0.000748* | 50.0 (13.9)  6.7 (1.3)  p=0.000937* | 75.2 (32.6) | 2.2 (1.2) |
| MARD /MARD (266) | **28.3 (5.1)** | **67.0 (9.0)** | **42.0 (8.0)**  **6.0 (0.7)** | **72.9 (30.7)** | **1.7 (1.1)** |
| /SIDD (5) | 28.7 (4.3) | 56.3 (7.9)  p=0.023197* | 49.0 (3.0)  6.6 (0.3)  p=0.030004 * | 63.3 (48.0) | 1.8 (3.1) |
| /SIRD (10) | 29.6 (9.3) | 68.9 (4.2) | 43.2 (8.5)  6.1 (0.8) | 101.2 (32.8)  p=0.023373* | 2.6 (0.7)  p=0.000537* |
| /MOD (10) | 31.0 (6.8) | 56.5 (17.3)  p=0.012895* | 42.0 (5.8)  6.0 (0.5) | 73.5 (28.7) | 2.0 (1.2) |

Data are median (IQR), † all values reported when n<3, ‡ minimum and maximum values reported, when n=3. SAID/SIDD/SIRD, Severe autoimmune/insulin-deficient/insulin-resistant diabetes; MOD/MARD, mild obesity-/age-related diabetes; BMI, body mass index; HbA1c, hemoglobin A1c; HOMA2-B, Homeostasis model assessment of β-cell function; HOMA2-IR, Homeostasis model assessment of insulin resistance. *Bonferroni-corrected p-values (Kruskal-Wallis) for the difference compared with those who were allocated to the same subgroup at both time-points (SIDD/SIDD, SIRD/SIRD, MOD/MOD, MARD/MARD).

**Table S8. Change in the variables used for the cluster-based subgrouping between registration and follow-up stratified according to the cluster subgroup assigned at registration and at follow-up (first column).**

| Cluster subgroup at registration /follow-up (n) | Change from registration to follow-up | | | |
| --- | --- | --- | --- | --- |
|  | **BMI (kg/m^2^)** | **HbA1c (mmol/mol)**  **(%)** | **HOMA2-B** | **HOMA2-IR** |
| SIDD /SIDD (2) | **-1.3 (-)** | **-9.1 (-)**  **-0.8 (-)** | **+0.3 (-)** | **-0.1 (-)** |
| /SIRD (1) | -0.6 (-) | -44.0 (-)  -6.2 (-) | -12.4 (-) | +3.3 (-) |
| /MOD (5) | +1.8 (2.4) | -32.0 (26.0)  -2.9 (2.4) | +28.0 (34.8) | -0.3 (2.1) |
| /MARD (8) | +1.8 (2.2) | -50.2 (20.0)  -4.6 (1.8) | +21.4 (76.1) | -0.3 (2.1) |
| SIRD /SIRD (11) | **-0.3 (3.4)** | **+2.0 (9.0)**  **+0.2 (0.8)** | **+5.0 (55.2)** | **+0.1 (2.9)** |
| /MOD (15) | 0.0 (4.0) | +2.0 (11.0)  +0.2 (1.0) | -59.2 (50.3)  p=0.000885* | -3.0 (4.9)  p=0.001717* |
| /MARD (26) | -1.0 (2.9) | -0.5 (11.5)  -0.0 (1.0) | -58.6 (69.4)  p=0.002433* | -1.9 (3.3)  p=0.010269* |
| MOD /MOD (113) | **-1.1 (3.6)** | **+1.5 (9.0)**  **+0.1 (0.8)** | **-5.0 (38.0)** | **-0.1 (1.0)** |
| /SIDD (3) | -1.4 (-) | +39.0 (-)  +3.6 (-) | -56.8 (-) | +0.3 (-) |
| /SIRD (3) | -1.4 (-) | -4.0 (-)  -0.4 (-) | +60.3 (-)  p=0.027407* | +0.7 (-) |
| /MARD (29) | -4.3 (3.9)  p=0.000002* | -3.3 (9.4)  -0.3 (0.9)  p=0.011099* | +0.3 (20.2) | -0.5 (1.2)  p=0.013146* |
| MARD /MARD (266) | **-0.8 (1.9)** | **+2.0 (6.0)**  **+0.2 (0.6)** | **-0.8 (31.2)** | **0.0 (0.9)** |
| /SIDD (5) | -0.8 (2.8) | +26.0 (12.0)  +2.4 (1.1)  p=0.000558* | -30.6 (50.7) | +0.5 (2.7) |
| /SIRD (10) | +0.3 (2.0) | +3.5 (8.0)  +0.3 (0.7) | +48.2 (55.8)  p=0.000117* | +1.8 (2.3)  p=0.000011* |
| /MOD (10) | +1.9 (3.3)  p=0.000147* | +8.0 (10.3)  +0.7 (0.9)  p=0.020375* | -4.5 (25.9) | +0.1 (0.5) |

Data are shown as median (IQR) SAID/SIDD/SIRD, Severe autoimmune/insulin-deficient/insulin-resistant diabetes; MOD/MARD, mild obesity-/age-related diabetes; BMI, body mass index; HbA1c, hemoglobin A1c; HOMA2-B, Homeostasis model assessment of β-cell function; HOMA2-IR, Homeostasis model assessment of insulin resistance. *Bonferroni-corrected p-values (Kruskal-Wallis test) are reported against subgroups reallocated to original subgroup (SIDD to SIDD, SIRD to SIRD, MOD to MOD, MARD to MARD).

**Table S9. Logistic regression: Odds of liver fibrosis (LSM ≥ 8kPa vs < 5kPa) based on variables measured at registration.**

|  | **Unadjusted** | | **Adjusted for BMI at registration** | |
| --- | --- | --- | --- | --- |
| **Variables** | **OR**  **(95% CI)** | **P** | **OR**  **(95% CI)** | **P** |
| Age | 1.02  (0.98-1.05) | 0.430 | 1.03  (0.99-1.07) | 0.198 |
| BMI_reg_ (kg/m2) | 1.09  (1.01-1.17) | 0.020 | - | - |
| HOMA2-IR_reg_ | 1.02  (0.81-1.27) | 0.884 | 1.00  (0.80-1.25) | 0.998 |
| HOMA2-IR_reg_ (INS_endo+exo_) * | 0.95  (0.76-1.18) | 0.619 | 0.95  (0.76-1.18) | 0.618 |
| HbA1c_reg_ (mmol/l) | 1.00  (0.98-1.03) | 0.625 | 1.01  (0.99-1.03) | 0.388 |
| Adipo-IR_reg_ (mmol/l x μU/ml) * | 1.03  (0.99-1.09) | 0.216 | 1.03  (0.97-1.08) | 0.368 |
| SAID ** | - | - | - | - |
| SIDD | 6.00  (0.48-75.34) | 0.165 | 5.58  (0.38-81.67) | 0.209 |
| SIRD | 6.50  (1.09-38.63) | 0.040 | 4.80  (0.69-33.47) | 0.114 |
| MOD | 2.85  (0.65-12.51) | 0.165 | 0.59  (0.09-3.87) | 0.583 |
| MARD | 1.70  (0.40-7.20) | 0.471 | 2.19  (1.10-1.41) | 0.341 |

A sensitivity analysis was conducted excluding those on insulin (n=83) with similar results than in the overall analysis. BMI, body mass index; HOMA2-IR, Homeostasis model assessment of insulin resistance; insulin concentration was analyzed with two methods, one mainly detecting endogenous insulin (INS_endo_ ), the other detecting both endogenous insulin and insulin analogues (INS_endo+exo_); HbA1c, haemoglobin A1c; Adipo-IR, Adipose tissue insulin resistance -index;* LSM ≥ 8kPa, n=39, LSM < 5kPa, n=15, **The SAID group was used as a reference for the other subgroups.

**Table S10. Univariate linear regression analysis of the association between liver steatosis (CAP, continuous attenuation parameter) at the study visit and different variables measured at the study visit (shown with and without those on insulin) or at registration (N=176).**

|  | Study visit | | | Study visit (no users of insulin) | | | Registration | |
| --- | --- | --- | --- | --- | --- | --- | --- | --- |
| Variable | **β-coefficient** | **p-value** | **β-coefficient** | | **p-value** | **β-coefficient** | | **p-value** |
| BMI | **0.462** | **<0.001** | **0.493** | | **<0.001** | **0.416** | | **<0.001** |
| K_ITT_ | -0.105 | 0.191 | -0.100 | | 0.250 | - | | - |
| fS-Triglycerides | **0.383** | **<0.001** | **0.345** | | **<0.001** | - | | - |
| fS-FFA* | **0.171** | **0.024** | **0.205** | | **0.012** | 0.093 | | 0.342 |
| HOMA2-IR | **0.268** | **<0.001** | **0.397** | | **<0.001** | **0.216** | | **0.024** |
| HOMA2-IR (INS_endo+exo_)* | **0.233** | **0.002** | **0.445** | | **<0.001** | 0.131 | | 0.188 |
| Adipo-IR* | **0.347** | **<0.001** | **0.397** | | **<0.001** | **0.302** | | **0.002** |
| Adipo-IR (INS_endo+exo_)* | **0.257** | **<0.001** | **0.460** | | **<0.001** | **0.265** | | **0.007** |
| Age | -0.121 | 0.111 | **-0.254** | | **0.002** | -0.100 | | 0.185 |
| B-HbA1c | 0.112 | 0.138 | **0.184** | | **0.025** | 0.042 | | 0.576 |
| fS-Insulin_endo_* | **0.319** | **<0.001** | **0.356** | | **<0.001** | **0.227** | | **0.019** |
| fP-Glucagon* | **0.267** | **<0.001** | **0.262** | | **0.001** | 0.137 | | 0.187 |
| Sex | **-0.149** | **0.048** | -0.071 | | 0.386 | **-0.149** | | **0.048** |

Abbreviations: BMI, body mass index; K_ITT_, first-order rate constant for glucose disappearance; LSM, Liver stiffness measurement; FFA, Free fatty acid; HOMA2-IR, Homeostasis model assessment of insulin resistance; insulin concentration was analyzed with two methods, one mainly detecting endogenous insulin (INS_endo_ ), the other detecting both endogenous insulin and insulin analogues (INS_endo+exo_); Adipo-IR, Adipose tissue insulin resistance -index; HbA1c, haemoglobin A1c. *N=119 patients at registration.

**Table S11. Univariate linear regression analysis of the association between liver fibrosis (LSM, liver stiffness measurement) at the study visit and different variables measured at the study visit (shown with and without those on insulin) or at registration (N=176).**

|  | Study visit | | Study visit/ exogenous insulin-users excluded | | Registration | |
| --- | --- | --- | --- | --- | --- | --- |
| Variable | **β-coefficient** | **p-value** | **β-coefficient** | **p-value** | **β-coefficient** | **p-value** |
| BMI | **0.341** | **<0.001** | **0.339** | **<0.001** | **0.257** | **<0.001** |
| K_ITT_ | **-0.203** | **0.011** | **-0.214** | **0.013** | - | - |
| Triglycerides | **0.156** | **0.039** | **0.174** | **0.034** | - | - |
| CAP | **0.310** | **<0.001** | **0.292** | **<0.001** | - | - |
| FFA | **0.213** | **0.004** | **0.190** | **0.020** | 0.063 | 0.516 |
| HOMA2-IR | **0.296** | **<0.001** | **0.448** | **<0.001** | 0.155 | 0.107 |
| HOMA2-IR (INS_endo+exo_)* | **0.269** | **<0.001** | **0.433** | **<0.001** | 0.146 | 0.140 |
| Adipo-IR* | **0.322** | **<0.001** | **0.429** | **<0.001** | **0.282** | **0.003** |
| Adipo-IR (INS_endo+exo_)* | **0.282** | **<0.001** | **0.434** | **<0.001** | **0.200** | **0.042** |
| Age | 0.108 | 0.154 | 0.050 | 0.542 | 0.099 | 0.191 |
| HbA1c | 0.107 | 0.159 | 0.124 | 0.133 | 0.017 | 0.824 |
| fS-Insulin_endo_ | **0.276** | **<0.001** | **0.402** | **<0.001** | **0.228** | **0.018** |
| Glucagon* | **0.198** | **0.009** | **0.194** | **0.018** | 0.048 | 0.643 |
| Sex | 0.024 | 0.753 | 0.046 | 0.577 | 0.024 | 0.753 |

Abbreviations: BMI, body mass index; K_ITT_, first-order rate constant for glucose disappearance; CAP, continuous attenuation parameter; FFA, Free fatty acid; HOMA2-IR, Homeostasis model assessment of insulin resistance; insulin concentration was analyzed with two methods, one mainly detecting endogenous insulin (INS_endo_ ), the other detecting both endogenous insulin and insulin analogues (INS_endo+exo_); Adipo-IR, Adipose tissue insulin resistance -index; HbA1c, hemoglobin A1c. *N=119 patients at registration (21 SAID, 6 SIDD, 23 SIRD, 28 MOD, 41 MARD).

**Methods – liver steatosis & alcohol-intake**

Six (2 SIRD, 2 MOD, 2 MARD) patients consumed more than 23/15 (men/women) doses (12g) of alcohol per week (categorized as excess alcohol consumption). Their removal from the analysis did not significantly influence the results and the self-reported amount of alcohol-use did not correlate with CAP (R=0.11, p=0.15) or LSM (R=-0.13, p=0.09).
